# Classifying progression status statements from radiology exams among non-small cell lung cancer patients using natural language processing

**DOI:** 10.1101/2021.11.20.21266642

**Authors:** Anahita Davoudi, Shun Yu, Abigail Doucette, Peter Gabriel, Mark Miller, Heather Williams, Heena Desai, Anh Le, Christian J Stoeckert, Kara Maxwell, Danielle L. Mowery

## Abstract

Although NLP has been used to support cancer research more broadly, the development of NLP algorithms to extract evidence of progression from clinical notes to support lung cancer research is still in its infancy. In this study, we trained supervised machine learning classifiers using rich semantic features to detect and classify statements of progression status from radiology exams. Our progression status classifier achieves high F1-scores for detecting and discerning progression (0.80), stable (0.82), and not relevant (0.92) sentences, demonstrating promising performance. We are actively integrating these extractions with structured electronic health record data using ontologies to instantiate a longitudinal model of progression among non-small cell lung cancer patients.

## Introduction

The prediction of patient outcomes in oncology relies on factors including pathological features, performance status, comorbidities, and a variety of tumor molecular alterations. Inherited genetic factors may also play a role in prediction of recurrence and overall survival in cancer patients. Both rare and common genetic variation is associated with cancer risk and outcome; however, the contribution of individual common variants or single nucleotide polymorphisms (SNPs) is small. Weighted polygenic risk scores (PRS) combining multiple SNPs are associated with risk in many cancers; however, it is unknown if PRS may be associated with outcomes in cancer. To study this hypothesis, robust and high throughput methods for determining a patient’s lung cancer outcomes with regards to diagnosis, time-to-progression, recurrence, and survival are needed.^1^

Technology companies and cancer registries have developed services in which trained oncology experts manually abstract progression status and other related information from the clinical notes of oncology patients’ electronic health records. However, the ascertainment of this data can be delayed several months behind the time of real-world data generated at the point of care. Natural language processing (NLP) is a technology at the intersection of biomedical informatics, computational linguistics, computer science, and artificial intelligence.^2, 3^ NLP can expedite the extraction of study variables i.e., progression status and derivation of cancer outcomes from clinical notes for more timely, scalable, and cost-effective to support use cases including but not limited to clinical trial matching, genotype/phenotype studies, pharmacoepidemiology, and cancer registries.^4–6^

### Leveraging natural language processing for lung cancer research

Although NLP has been used to support cancer research more broadly,^5–10^ the development of NLP algorithms to extract evidence of progression from clinical notes to support lung cancer research is still in its infancy. For example, Krishner et al. trained a logistic regression model using metastatic terms related to progression in bladder cancer, melanoma, non-small cell lung cancer (NSCLC), breast cancer, colorectal cancer, prostate cancer, and renal cell carcinoma. They classified patients according to cancer type and status: metastatic highly likely, metastatic likely, unknown, non-metastatic likely, and non-metastatic highly unlikely.^4^ For categorizing NSCLC patients, their classifier achieved reasonable precision of 87.3% and recall of 84.0%. Kehl et al. trained a convolutional neural network to predict whether the text within the assessment and plan section of radiology exams indicate cancer present, cancer progressing/worsening, and cancer responding/improving.^7^ On their test set, the progression classifier achieved a best F1-score of 62% for progression/worsening and F1-score of 64% for response/improvement. Although these results are promising, the Krishner et al. classifier results are not reported by subtype, i.e., metastatic likely or progression status making it hard to determine possible challenges in classifying the metastatic classes and comparing to our work. Furthermore, the Kehl et al. classifier performs moderately across both classes. Our study expands beyond these related works with two main objectives: 1) classifying diagnostic statements about progression status using deep learning and rich semantic features and 2) learning the informativeness of deep semantic features for classifying progression status.

## Method

In this University of Pennsylvania Institute Review Board-approved pilot study (#834430), we applied a transfer learning approach to predict progression status derived from data stored in two institutional databases, Flatiron and the Oncology Research and Quality Improvement Datamart. The Flatiron database contains real-world clinical data collected from the electronic health records (EHRs) of academic health systems documented by cancer care providers throughout the United States. These longitudinal care data are reviewed by a team of oncology abstractors that extract cancer outcomes including stage, progression-free survival, mortality among other outcomes. The Oncology Research and Quality Improvement Datamart (ORQID) is a data warehouse that aggregates together structured data extracted from PennChart (Penn Medicine’s EHR), the Penn Medicine Cancer Registry (accredited by the American College of Surgeons Commission on Cancer), the ARIA® Radiation Oncology Information System, the Penn Center for Personalized Diagnostics genomic variant database, and several other clinical systems. We queried all stage 3+ non-small cell lung cancer patients identified in both the Flatiron database and ORQID (n=4813 patients) and their associated radiology notes.

### Annotating sentences about progression status using expert opinion

We randomly selected 100 patients and their associated CT chest exams from this dataset. Clinical oncologist (co-author, SY) annotated each sentence from the impression sections of the associated computerized tomography chest exams into one of the following progression status classes:

- **Progression**, mentions of increased size of solid masses or spread of cellular cancer
- **Stable**, mentions of solid masses without interval change or lack of cellular cancer spread

This resulted in 1,270 sentences annotated as either progression or stable. Sentences that could not be classified into these groups were automatically encoded as **Not relevant**. Progression can occur locally in the lungs and spread to other areas of the body e.g., brain, bone, adrenal glands, etc. To increase the number of training cases of representing progression or metastasis across other cancer types, we applied an unsupervised, deep learning approach of sampling sentences from any radiology note types using Bio+Clinical Bidirectional Encoder Representations from Transformers (BERT) and cosine similarity of 0.95-0.99.^11^ These sentences were validated by SY adding 194 sentences to the corpus. We randomly selected 1,821 not relevant sentences (also not annotated) from the 100 patients resulting in a total of 3,285 sentences for classifying progression status.

### Classifying sentences by progression status

For sentence-level classification of progression status -- **progression, stable**, or **not relevant** -- we randomly split the annotated dataset into training (80%; n=2,628 sentences) and testing (20%; n=657 sentences) sets. We trained and tested our progression status classifier using four types of features:

- **Corpus N-Grams (CN):** From the training set, we generated n-grams using word windows of n=1-3. We applied term-frequency/inverse document frequency (TF-IDF) to identify the most informative terms throughout the corpus.
- **Radiology Entities (RE):** We encoded named entities using models from STANZA.^12^ From the radiology NER model (radiology package), we encoded 5 types of entities: *anatomy, observation, anatomy modifier, observation modifier*, and *uncertainty*. From the i2b2 NET model (i2b2 package), we encoded *problems, treatments*, and *tests*. From the Cancer NER model (bionlp13cg package), we encoded 16 entity types in cancer entities: *amino acid, anatomical system, cancer, cell, cellular component, developing anatomical structure, gene or gene product, immaterial anatomical entity, multi-tissue structure, organ, organism, organism subdivision, organism substance, pathological formation, simple chemical, tissue*.
- **Contextual Terms (CT):** We generated contextual features to capture negation and uncertainty, non-patient experiencer, and historicity based on the pyConTextNLP lexicon.^8, 13^ For example, entities representing uncertainty include: *definite existence, probable existence, possible existence, ambivalent existence, possible negated existence, probable negated existence*, and *definite negated existence*. Entities representing temporality include: *historical, acute*, and *future*. Other functional entities include: *pseudo negation* and *conjunction*.
- **Metastasis Terms (MT):** We generated features based on metastatic term lists from work by Krishner et al.^4^ in Appendix Table A1 of their manuscript. We included subtypes for *advanced bladder cancer, advanced melanoma, advanced NSCLC, metastatic breast cancer, metastatic colorectal cancer, metastatic prostate cancer*, and *metastatic renal cell carcinoma*.

Using these feature sets, we trained three machine learning (ML) algorithms: Logistic Regression, XGBoost, and LightGBM.

- **Logistic regression (LR)** learns a model to explain the relationship between all features and the class. We trained our model using an exhaustive grid search and L1-regularization to optimize performance.
- **Light gradient boosting machine (LGBM)** is a distributed high-performance gradient boosting model based on decision trees. This model has been found to produce better accuracy than other boosting algorithms by producing more complex trees leveraging a leaf-wise split approach.
- **Extreme gradient boosted trees (XGBoost)** learns a model to predict the residuals or errors of prior models while minimizing the loss of adding new models before combining models to make a final class prediction. Complex trees are generated applying level-wise split approach. The booster parameter was set to gblinear.

For our first experiment, we aimed to determine whether a deep learning model could outperform traditional supervised learning models using n-grams. We compared the best performing classifier with simple corpus n-grams (CN) to a deep learning, lexicon-based system trained using **Bio+Clinical Bidirectional Encoder Representations from Transformers (BERT)**. Bio+Clinical BERT is a BERT model that leverages pre-trained language representations initialized from PubMed article abstracts and PubMed Central article full texts and fine-tuned using a clinical corpus of notes (e.g., discharge summaries, physician notes, nursing notes, radiology reports, etc.) from the Medical Information Mart for Intensive Care (MIMIC version III) dataset.^11^ The Bio+Clinical BERT model is integrated with an XGBoost classifier in the final layer to classify sentences according to progression status.

For our second experiment, we aimed to determine the predictive value of including richer semantic features for predicting progression status. For each progression status class, we graphed the recall and precision curves of each ML classifier by additively including each feature group to show the added predictive power of each feature set to classification. Feature sets include: CN, CN+RE, CN+RE+CT, CN+RE+CT+MT. A workflow schematic can be found in **Figure 1**.

**Figure 1.**
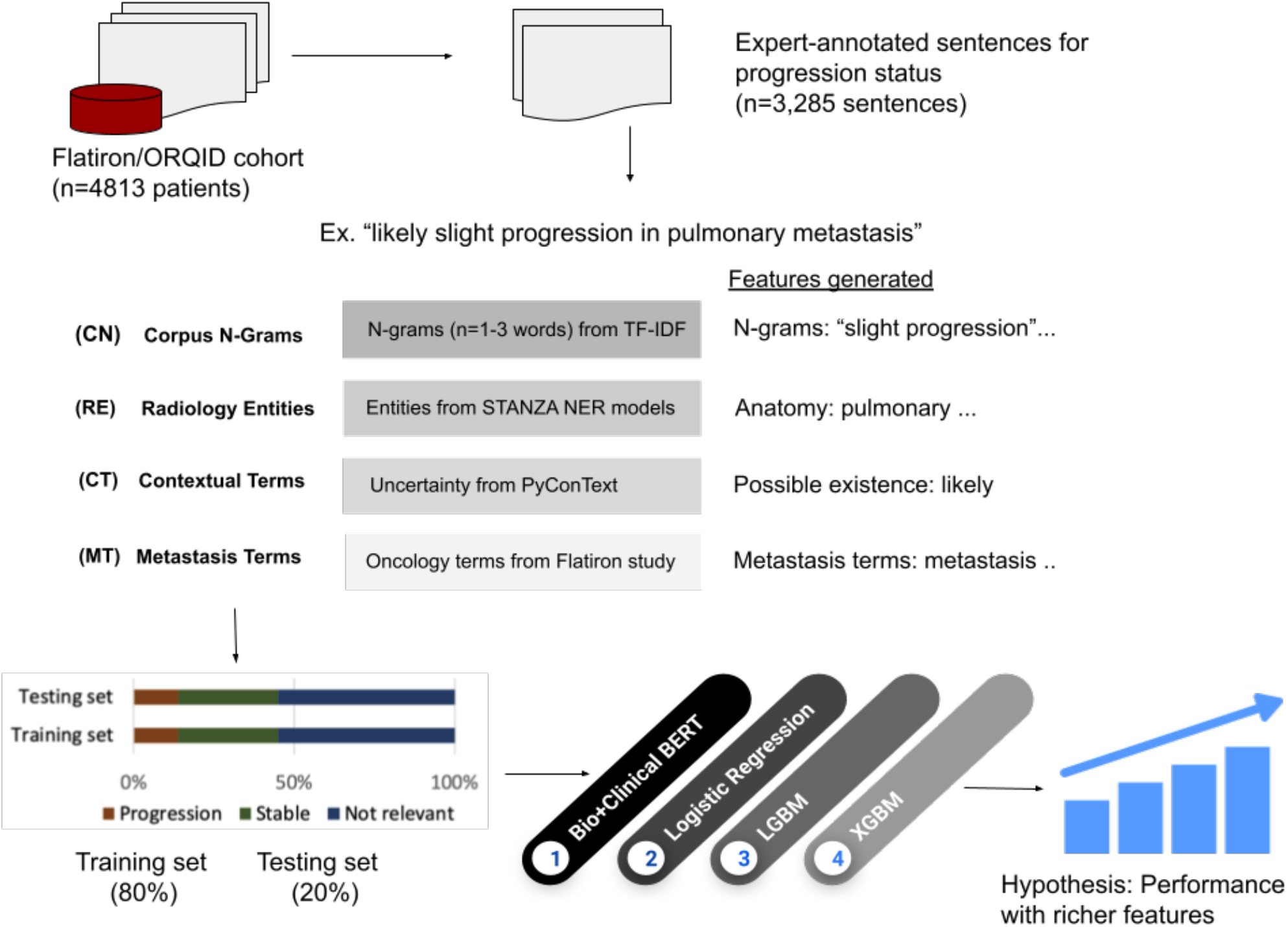
Workflow of annotation and automation of progression status.

## Results

In **Table 1**, the distribution across progression classes in the training and testing set were identical: not relevant (55%) followed by stable (31%) and progression (14%).

**Table 1.**
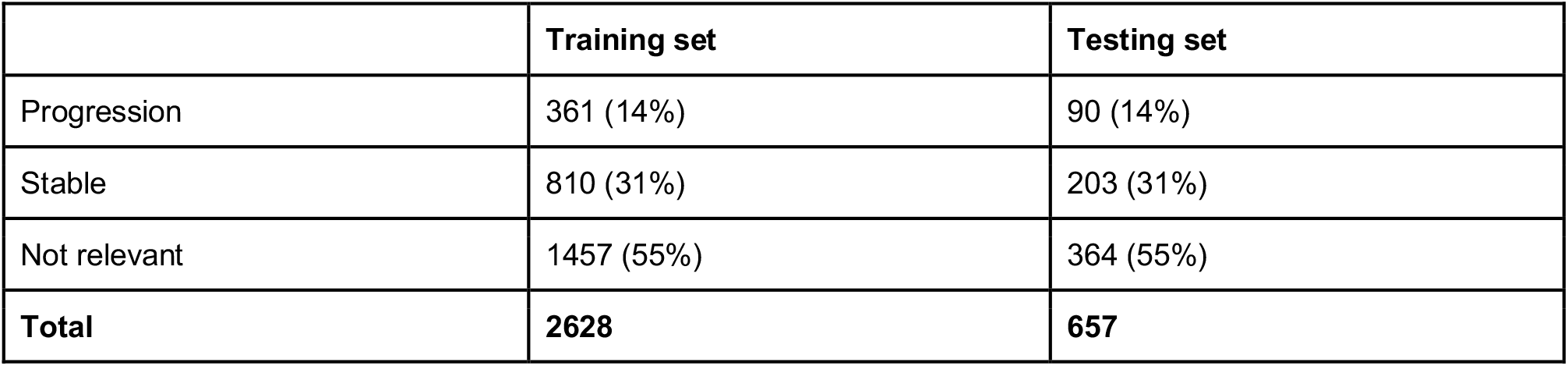
Annotated sentences characterized by training and testing sets.

In **Table 2**, we compare the performance of Bio+Clinical BERT+XGBM to simple corpus n-grams across classifiers. In terms of corpus n-grams, the highest performing classifier according to progression status class includes: progression (logistic regression; F1: 0.75), stable (LGBM; F1: 0.77), and not relevant (Bio+Clinical BERT+XGBM; F1: 0.92). We report the added predictive power when including richer semantic feature sets to train the non-BERT classifiers. For progression, the highest F1-score (0.80) was achieved leveraging corpus n-grams, radiology entities, contextual terms, and metastatic terms to train XGBoost. Similarly, for stable, the highest F1-score (0.82) was achieved with these same features to train logistic regression. For not relevant, the highest F1-score (0.92) was achieved using all semantic features except metastatic terms. For recall, by including richer semantic features, we observed up to 8 point increase for progression, 10 point increase for stable, and 2 point increase for not relevant. Similarly, for precision, we observed an up to 5 point increase for progression, 5 point increase for stable, and 3 point increase for not relevant.

**Table 2.** Mention-level classification of progression status for the test set only

|  | Embeddings |  |  | CN+RE |  |  | CN+RE+CT |  |  | CN+RE+CT+MT |  |  |
| --- | --- | --- | --- | --- | --- | --- | --- | --- | --- | --- | --- | --- |
| Bio+Clinical BERT + XGBM | P | R | F1 | P | R | F1 | P | R | F1 | P | R | F1 |
| Progression | 0.66 | 0.57 | 0.61 | -- | -- | -- | -- | -- | -- | -- | -- | -- |
| Stable | 0.75 | 0.74 | 0.75 | -- | -- | -- | -- | -- | -- | -- | -- | -- |
| Not relevant | **0.90** | **0.93** | **0.92** | -- | -- | -- | -- | -- | -- | -- | -- | -- |
| Macro average | 0.77 | 0.75 | 0.76 | -- | -- | -- | -- | -- | -- | -- | -- | -- |
| Micro average | 0.82 | 0.82 | 0.82 | -- | -- | -- | -- | -- | -- | -- | -- | -- |
|  | CN |  |  | CN+RE |  |  | CN+RE+CT |  |  | CN+RE+CT+MT |  |  |
| Logistic Regression | P | R | F1 | P | R | F1 | P | R | F1 | P | R | F1 |
| Progression | **0.75** | **0.76** | **0.75** | 0.73 | 0.73 | 0.73 | 0.75 | 0.74 | 0.75 | 0.71 | 0.83 | 0.77 |
| Stable | 0.79 | 0.74 | 0.77 | 0.81 | 0.78 | 0.79 | 0.79 | 0.76 | 0.78 | **0.81** | **0.84** | **0.82** |
| Not relevant | 0.89 | 0.93 | 0.91 | 0.91 | 0.92 | 0.91 | **0.91** | **0.93** | **0.92** | 0.89 | 0.92 | 0.90 |
| Macro average | 0.81 | 0.81 | 0.81 | 0.82 | 0.81 | 0.81 | 0.82 | 0.81 | 0.82 | 0.80 | 0.86 | 0.83 |
| Micro average | 0.84 | 0.85 | 0.84 | 0.85 | 0.85 | 0.85 | 0.85 | 0.85 | 0.85 | 0.84 | 0.88 | 0.86 |
|  | CN |  |  | CN+RE |  |  | CN+RE+CT |  |  | CN+RE+CT+MT |  |  |
| LGBM | P | R | F1 | P | R | F1 | P | R | F1 | P | R | F1 |
| Progression | 0.72 | 0.76 | 0.74 | 0.73 | 0.78 | 0.75 | 0.77 | 0.81 | 0.79 | 0.77 | 0.78 | 0.77 |
| Stable | **0.79** | **0.76** | **0.77** | 0.82 | 0.72 | 0.77 | 0.84 | 0.74 | 0.79 | 0.82 | 0.76 | 0.79 |
| Not relevant | 0.90 | 0.91 | 0.91 | 0.88 | 0.93 | 0.90 | 0.89 | 0.93 | 0.91 | 0.90 | 0.93 | 0.91 |
| Macro average | 0.80 | 0.81 | 0.81 | 0.81 | 0.81 | 0.81 | 0.83 | 0.83 | 0.83 | 0.83 | 0.82 | 0.82 |
| Micro average | 0.84 | 0.84 | 0.84 | 0.84 | 0.84 | 0.84 | 0.86 | 0.85 | 0.86 | 0.86 | 0.86 | 0.85 |
|  | **CN** |  |  | **CN+RE** |  |  | **CN+RE+CT** |  |  | **CN+RE+CT+MT** |  |  |
| **XGBoost** | **P** | **R** | **F1** | **P** | **R** | **F1** | **P** | **R** | **F1** | **P** | **R** | **F1** |
| Progression | 0.74 | 0.74 | 0.74 | 0.73 | 0.77 | 0.75 | 0.74 | 0.78 | 0.76 | **0.77** | **0.82** | **0.80** |
| Stable | 0.78 | 0.75 | 0.77 | 0.78 | 0.72 | 0.75 | 0.80 | 0.72 | 0.76 | 0.83 | 0.77 | 0.80 |
| Not relevant | 0.89 | 0.91 | 0.90 | 0.88 | 0.91 | 0.89 | 0.88 | 0.92 | 0.90 | 0.90 | 0.92 | 0.91 |
| Macro average | 0.80 | 0.80 | 0.80 | 0.80 | 0.80 | 0.80 | 0.81 | 0.81 | 0.81 | 0.83 | 0.84 | 0.84 |
| Micro average | 0.83 | 0.84 | 0.84 | 0.83 | 0.83 | 0.83 | 0.84 | 0.84 | 0.84 | 0.86 | 0.86 | 0.86 |

## Discussion

In this pilot study, we aimed to classify diagnostic statements about progression status using deep learning and rich semantic features and to learn the predictive value of deep semantic features for classifying progression status. We learned that the Bio+Clinical BERT model could accurately discern not relevant sentences with high performance, but could not detect and discern progression (XGBoost) and stable (LGBM) statements better than the other supervised classifiers. In this case, an ensemble approach could boost classification performance for relevant classes. We also learned that richer features could improve classification performance. We observe increases in precision and recall for progression and stable classes with richer feature sets compared to corpus n-grams alone.

In comparison to related works, our classifier achieved superior performance compared to Kehl et al. On their test set, the progression classifier achieved a best F1-score of 62% for progression/worsening and F1-score of 64% for response/improvement. Our XGBoost progression classifier achieved 18 points higher F1-score for progression; our logistic regression classifier achieved 18 points higher F1-score for stable. We hypothesize that rich negation/uncertainty and precision terms for metastasis resulted in notable improvements for progression and stable status because the classifiers were better able to detect progressions across non-NSCLC cancer types (bladder, breast, prostate, renal, etc.) and discern negations of progression (“tumor not increasing in size”) representing stable assertions. Unsurprisingly, metastasis terms (“mets”, “metastasis”) did not add more discriminatory power for not relevant sentences which largely don’t contain these terms. An error analysis suggests that our system could be improved with additional features. For example, some sentences were misclassified as not relevant rather than progression or stable due to absence of tumor size change descriptions (“smaller”) and inferences (“from 3.3 × 3.3 cm…previously measuring as 3.2 × 3.2 cm”). These cases will be handled by our commercial NLP system, Linguamatics, which has built-in libraries that support extraction of tumor size dimensions.

The implications of our system are important. Determination of progression-free survival stages heavily relies on evidence of progression or stable statements from radiology images. However, we understand that radiology imaging is one part of a larger clinical picture. A longitudinal model that integrates lines of therapy (LOT) changes from medications as well as progression statements and symptoms from clinical notes will be needed to infer progression and response by patients receiving treatment over time. We are actively encoding LOT and applying our progression status classifiers to generate additional features for instantiating an encounter-level, longitudinal model of progression using ontologies as part of the PennTURBO (Transforming and Unifying Research with Biomedical Ontologies)^14^ platform.

## Limitations and Future Work

Notably, we have only trained and tested our classifiers using Penn Medicine data and for NSCLC patients. However, we believe that the sublanguage used to describe progression status may generalize across academic medical centers and cancer types. In the future, we will assess the generalizability of our models in predicting progression status among gastric, pancreatic, and colon cancer patients from patient notes. We will also apply this classifier to the Penn Medicine Biobank lung cancer patient data to conduct genotype/phenotype association studies to better understand genetic risk and patterns of progression.

## Conclusion

Progression status mentions can be accurately extracted and encoded from radiology exams. This marks the first step toward a more comprehensive model of longitudinal progression to support cancer research.

## Data Availability

The classifiers produced in the present study will be openly available.

